# International Electronic Health Record-Derived COVID-19 Clinical Course Profiles: The 4CE Consortium

**DOI:** 10.1101/2020.04.13.20059691

**Authors:** Gabriel A Brat, Griffin M Weber, Nils Gehlenborg, Paul Avillach, Nathan P Palmer, Luca Chiovato, James Cimino, Lemuel R Waitman, Gilbert S Omenn, Alberto Malovini, Jason H Moore, Brett K Beaulieu-Jones, Valentina Tibollo, Shawn N Murphy, Sehi L’Yi, Mark S Keller, Riccardo Bellazzi, David A Hanauer, Arnaud Serret-Larmande, Alba Gutierrez-Sacristan, John J Holmes, Douglas S Bell, Kenneth D Mandl, Robert W Follett, Jeffrey G Klann, Douglas A Murad, Luigia Scudeller, Mauro Bucalo, Katie Kirchoff, Jean Craig, Jihad Obeid, Vianney Jouhet, Romain Griffier, Sebastien Cossin, Bertrand Moal, Lav P Patel, Antonio Bellasi, Hans U Prokosch, Detlef Kraska, Piotr Sliz, Amelia LM Tan, Kee Yuan Ngiam, Alberto Zambelli, Danielle L Mowery, Emily Schiver, Batsal Devkota, Robert L Bradford, Mohamad Daniar, APHP/Universities/INSERM COVID-19 research collaboration, Christel Daniel, Vincent Benoit, Romain Bey, Nicolas Paris, Patricia Serre, Nina Orlova, Julien Dubiel, Martin Hilka, Anne Sophie Jannot, Stéphane Bréant, Judith Leblanc, Nicolas Griffon, Anita Burgun, Melodie Bernaux, Arnaud Sandrin, Elisa Salamanca, Thomas Ganslandt, Tobias Gradinger, Julien Champ, Martin Boeker, Patricia Martel, Loic Esteve, Alexandre Gramfort, Olivier Grisel, Damien Leprovost, Thomas Moreau, Gael Varoquaux, Jill-Jenn Vie, Demian Wassermann, Arthur Mensch, Charlotte Caucheteux, Christian Haverkamp, Guillaume Lemaitre, Ian D. Krantz, Sylvie Cormont, Andrew M South, The Consortium for Clinical Characterization of COVID-19 by EHR (4CE), Tianxi Cai, Isaac S Kohane

## Abstract

We leveraged the largely untapped resource of electronic health record data to address critical clinical and epidemiological questions about Coronavirus Disease 2019 (COVID-19). To do this, we formed an international consortium (4CE) of 96 hospitals across 5 countries (www.covidclinical.net). Contributors utilized the Informatics for Integrating Biology and the Bedside (i2b2) or Observational Medical Outcomes Partnership (OMOP) platforms to map to a common data model. The group focused on comorbidities and temporal changes in key laboratory test values. Harmonized data were analyzed locally and converted to a shared aggregate form for rapid analysis and visualization of regional differences and global commonalities. Data covered 27,584 COVID-19 cases with 187,802 laboratory tests. Case counts and laboratory trajectories were concordant with existing literature. Laboratory tests at the time of diagnosis showed hospital-level differences equivalent to country-level variation across the consortium partners. Despite the limitations of decentralized data generation, we established a framework to capture the trajectory of COVID-19 disease in patients and their response to interventions.

## Introduction

The Coronavirus Disease 2019 (COVID-19) pandemic has caught the world off guard, reshaping ways of life, the economy, and healthcare delivery all over the globe. The virulence and transmissibility of responsible virus (SARS-CoV-2) is striking. Crucially, there remains a paucity of relevant clinical information to drive response at the clinical and population levels. Even in an information technology-dominated era, fundamental measurements to guide public health decision-making remain unclear. Knowledge still lags on incidence, prevalence, case-fatality rates, and clinical predictors of disease severity and outcomes. While some of the knowledge gaps relate to the need for further laboratory testing, data that should be widely available in electronic health records have not yet been effectively shared across clinical sites, with public health agencies, or with policy makers. At the time of this writing, more than three months after the earliest reports of the disease in China, only 5.8% of US cases reported to the CDC have clinical details included.^1^

Even before therapeutic trials are implemented, frontline clinicians are not yet benefitting from knowledge as basic as understanding the differences in the clinical course between male and female patients. ^2^ Through case studies and series, we have learned that COVID-19 can have multi-organ involvement. A growing literature has identified key markers of cardiac,^3^ immune,^4^ coagulation,^5^ muscle,^5,6^ hepatic,^7^ and renal^8^ injury and dysfunction, including extensive evidence of myocarditis and cardiac injury associated with severe disease. Laboratory perturbations in lactate dehydrogenase (LDH), C-reactive protein (CRP), and procalcitonin^9^ have been described. However, data from larger cohorts, linked to outcomes, remain unavailable.

Because electronic health records (EHRs) are not themselves agile analytic platforms, we have been successfully building upon the open source and free i2b2 (for Informatics for Integrating Biology and the Bedside) toolkit^10-17^ to manage, compute, and share data extracted from EHRs. In response to COVID-19, we have organized a global community of researchers, most of whom are or have been members of the i2b2 Academic Users Group, to rapidly set up an *ad hoc* network that can begin to answer some of the clinical and epidemiological questions around COVID-19 through data harmonization, analytics, and visualizations. The Consortium for Clinical Characterization of COVID-19 by EHR (4CE)—pronounced “foresee”—comprises partner hospitals from five countries.

Our early efforts aim to consolidate, share, and interpret data about the clinical trajectories of the infection in patients with a first focus on laboratory values and comorbidities. This initial report seeks (a) to establish the accessibility and suitability of data from electronic medical record for COVID-19 patients; (b) to learn about the clinical trajectories of patients; (c) to facilitate evaluation and communication of the utility of various laboratory tests and therapies; and (d) to contribute data, reproducible data mining and visualization workflows, and learnings to a global network and the broader public.

Here, we report on initial results and the structure of a new, rapidly formed network designed to be a highly scalable system, now implemented at 23 sites. The international scope of our collaboration allows us to identify some of the similarities in clinical course and a few country-specific variations. We recognize that these early data are incomplete and are subject to many biases and limitations, which constrain the conclusions we can currently draw. However, we believe the sources of our data and the mechanism we have established for sharing them are sound, reproducible, and scalable. We also hope our results to-date will encourage other sites to share data and contribute to this important research effort.

## Methods

### Selection of Laboratory Values

Multiple studies have reported significant abnormalities in several laboratory tests in patients with COVID-19. Studies have shown abnormalities in cardiac, hepatic, renal, immune, and coagulation physiology. Those laboratory results are associated with both disease presentation and severity. For this initial study, we selected a subset of laboratories that are commonly performed, as identified by the Regenstrief Institute responsible for the Logical Objects, Identifiers, Names and Codes (LOINC) standard,^18^ and had been previously associated with worse outcomes in COVID-19 patients. Based on the meta-analysis of Lippi and Plebani,^19^ we focused on 14 laboratory studies that are commonly performed: alanine aminotransferase (ALT), aspartate aminotransferase (AST), total bilirubin (Tbili), albumin, cardiac troponin (high sensitivity), lactate dehydrogenase (LDH), D-dimer, white blood cell count (WBC), lymphocyte count, neutrophil count, procalcitonin, and prothrombin time. LOINC codes were identified for each laboratory study as well as the units and reference ranges.

### Cohort Identification

All patients who received a polymerase chain reaction (PCR) confirmed diagnosis of COVID-19 were included in the data collection. Some hospitals only included patients who were admitted to the hospital while others included all patients for whom the test was positive.

### Data Collection and Aggregation

Sites obtained the data for their files in several ways. Most sites leveraged the open source i2b2 software platform already installed at their institution^20^ which supports query and analysis of clinical and genomics data. More than 200 organizations worldwide use i2b2 for a variety of purposes, including identifying patients for clinical trials, drug safety monitoring, and epidemiology research. Most 4CE sites with i2b2 used database scripts to directly query their i2b2 repository to calculate counts needed for data files. Institutions without i2b2 used their own clinical data warehouse solutions and querying tools to create the files. In some cases, a hybrid method was used that leveraged different data warehouse platforms to fill in i2b2 gaps. For example, Assistance Publique – Hôpitaux de Paris (APHP), the largest hospital system in Europe, aggregates all EHR data from 39 hospitals in Paris and its surroundings. APHP exported data from the Observational Medical Outcomes Partnership (OMOP) Common Data Model for transformation to the shared format.

Each site generated four data tables, saved as comma-separated values (CSV) files. To protect patient privacy, the files we report contain only aggregate counts (no data on individual patients). In order to further protect patient identity, small counts were obfuscated (see below), since an aggregate count of “1” represents an individual patient. By computing these values locally and only sharing the aggregate data, sites were able to obtain institutional approval more rapidly.

The first file, *DailyCounts.csv*, contained one row per calendar date. Each row included the date, the number of new COVID-19 patients, the number of COVID-19 patients in an intensive care unit (ICU), and the number of new deaths from COVID-19.

The second file, *Demographics.csv*, contained counts of the total number of COVID-19 patients, broken down by sex and age group (0-2, 3-5, 6-11, 12-17, 18-25, 26-49, 50-69, 70-79, and 80+ years old).

The third file, *Labs.csv*, described the daily trajectories of select laboratory tests. Each row corresponded to a laboratory test (identified using a LOINC code) and the number of days since a patient had a positive COVID-19 test, ranging from −6 (one week before the test result) to 1 (the day of the test result) to N (the day the file was created). The values in each row are the number of patients who have a test result on that day and the mean and standard deviation of the test results.

The fourth file, *Diagnoses.csv*, lists all the diagnoses recorded in the EHR for COVID-19 patients, starting from one week before their positive COVID-19 test to the present, with the count of the number of patients with the corresponding ICD-9 or ICD-10 code.

Sites optionally obfuscated the values in any of these files by replacing small counts with “-1”. Sites indicated missing data or data that they were unable to obtain (e.g., whether patients were in an ICU) with “-2”.

Sites uploaded their files to a private shared folder. These files were merged into four combined files that included totals from individual sites. Each value in the combined file had four components: (1) number of sites with unmasked values; (2) sum of those values; (3) number of sites with obfuscated values; and (4) sum of the obfuscation thresholds for those sites. For example, if five sites reported values 25, 15, −1 (between 00 and 9 patients), −1 (between 00 and 4 patients), −1 (between 00 and 4 patients), then the combined file listed two unmasked sites with a total of 40 patients and three masked sites with up to 9+4+4=17 patients. From this, it was inferred that there were between 40 and 57 patients. Given the large geographic distance between our sites, we assumed that each COVID-19 patient was only represented in one EHR. The combined *Labs.csv* file contained a weighted average (rather than the sum) of the unmasked mean test results from each site.

### ICD Mapping

Diagnosis codes were submitted from the sites as either international clinical diagnosis (ICD)-9 or ICD-10 billing codes. ICD-9 diagnosis codes were mapped to ICD-10 by first attempting to match the ICD-9 codes to child concepts of ICD-10 codes in the Accrual to Clinical Trials (ACT) ICD-10➔ICD-9 ontology.^21^ In the cases where no match was found in the ACT ontology, ICD-9 codes were matched to the ICD-10 codes that shared a common concept unique identifier (CUI) in the 2019 build of the US National Library of Medicine’s (NLM’s) Unified Medical Language System (UMLS).^22^

### Data Sharing and Visualization

We created a website hosted at https://covidclinical.net to provide interactive visualizations of our datasets as well as direct access to all shareable data collected for this publication. Data aggregation and publication processes are shown in Figure 1. Visualizations were implemented using Python and Altair (http://altair-viz.github.io/) in Jupyter Notebooks (https://jupyter.org), all of which are freely available on the website. The Vega visualizations (http://vega.github.io) generated by Altair were embedded into a Jekyll-based site (http://jekyllrb.com/) that was hosted on Amazon Web Services.

**Figure 1.**
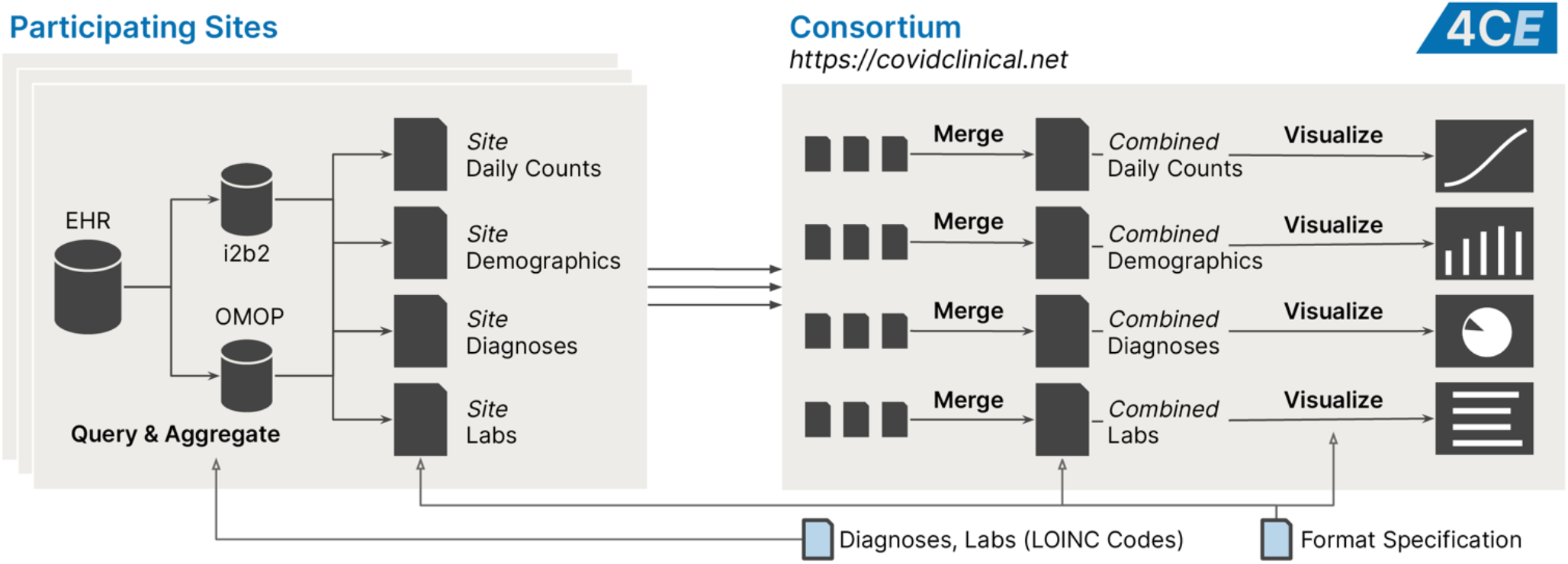
Overview of data collection and analysis.

### Data Availability

Data files for daily counts, demographics, diagnosis, and labs datasets are available at https://covidclinical.net.

### Informed Consent/IRB Statement

Each institution reported obtaining proper institutional review board approval for data sharing. Certifications of waivers or approval were collected by the consortium. As data were transmitted in aggregate, no patient level data were available from any site.

## Results

### Demographic and Consortium Level Data

Over a span of three weeks, 96 total hospitals in the US (45), France (42), Italy (5), Germany (3), and Singapore (1) contributed data to the consortium. This was represented by 23 data collaboratives across these five countries. A total of 27,584 patients with COVID-19 diagnosis were included in the dataset, with data covering January 1, 2020 through April 11, 2020. We collected 187,802 laboratory values and harmonized them across sites. Thirteen percent of sites submitted complete data sets that included values for each laboratory (39.1% for at least 13, and 43.5% for at least 12 of the 14 laboratory measurements). Breakdown of country level data is shown in Table 1.

**Table 1.** Sites contributing data to the consortium.

| Healthcare System | Acronym | City | Country | Population | Hospitals | Beds | Inpatient discharges/year |
| --- | --- | --- | --- | --- | --- | --- | --- |
| Assistance Publique - Hôpitaux de Paris | APHP | Paris | France | Adult & Pediatric | 39 | 20,098 | 1,375,538 |
| Bordeaux University Hospital | FRBDX | Bordeaux | France | Adult & Pediatric | 3 | 2,676 | 130,033 |
| Erlangen University Hospital | UKER | Erlangen | Germany | Adult & Pediatric | 1 | 1,400 | 65000 |
| Medical Center, University of Freiburg | UKFR | Freiburg | Germany | Adult & Pediatric | 1 | 1,660 | 71,500 |
| University Medicine Mannheim | UMM | Mannheim | Germany | Adult & Pediatric | 1 | 1,352 | 50,748 |
| ICSM Pavia Hospital | ICSM1 | Pavia | Italy | Adult | 1 | 426 | 8,616 |
| ICSM Lumezzane/Brescia Hospitals | ICSM5 | Lumezzane/<br>Brescia | Italy | Adult | 1 | 149 | 1,296 |
| ICSM Milano Hospital | ICSM20 | Milan | Italy | Adult | 1 | 200 | 2,432 |
| Policlinico di Milano | POLIMI | Milan | Italy | Adult &<br>Pediatric | 1 | 900 | 40,000 |
| ASST Papa Giovanni XXIII<br>Bergamo | HPG23 | Bergamo | Italy | Adult &<br>Pediatric | 1 | 1,080 | 45,000 |
| National University Hospital | NUH | Singapore | Singapore | Adult &<br>Pediatric | 1 | 1,556 | 100,977 |
| Boston Children's Hospital | BCH | Boston, MA | USA | Pediatric | 1 | 404 | 28,000 |
| Beth Israel Deaconess Medical<br>Center | BIDMC | Boston, MA | USA | Adult | 1 | 673 | 40,752 |
| Children's Hospital of Philadelphia | CHOP | Philadelphia,<br>PA | USA | Pediatric | 1 | 564 | 25,406 |
| University of Kansas Medical Center | KUMC | Kansas City,<br>KS | USA | Adult &<br>Pediatric | 1 | 794 | 54,659 |
| Mayo Clinic | MAYOC | Rochester,<br>MN | USA | Adult &<br>Pediatric | 1 | 2,059 | 100,000 |
| Mass General Brigham (Partners<br>Healthcare) | MGB | Boston, MA | USA | Adult &<br>Pediatric | 10 | 3,418 | 163,521 |
| Medical University of South<br>Carolina | MUSC | Charleston,<br>SC | USA | Adult &<br>Pediatric | 8 | 1,600 | 55,664 |
| University of Pennsylvania | UPenn | Philadelphia,<br>PA | USA | Adult | 5 | 2,469 | 118,188 |
| University of California, LA | UCLA | Los Angeles,<br>CA | USA | Adult &<br>Pediatric | 2 | 786 | 40,526 |
| University of Michigan | UMICH | Ann Arbor,<br>MI | USA | Adult &<br>Pediatric | 3 | 1,000 | 49,008 |
| University of North Carolina at<br>Chapel Hill | UNC | Chapel Hill,<br>NC | USA | Adult &<br>Pediatric | 11 | 3,095 | 52,000 |
| UT Southwestern Medical Center | UTSW | Dallas, TX | USA | Adult | 1 | 608 | 26,905 |
|  |  |  |  | **Total** | **96** | **45,352** | **2,444,792** |

Demographic breakdown by age and sex is shown in Figure 2. Age distribution was different across countries and consistent with previously identified patterns. In particular, patients from Italy were more commonly over the age of 70 relative to other countries.^23^ US institutions, despite representing a large number of active infections, had the lowest percentage of elderly patients diagnosed with COVID-19. Germany, with its three included hospitals and relatively small number of patients, was more similar to the US and had an increased number of male patients in the 50-59 age group.

**Figure 2.**
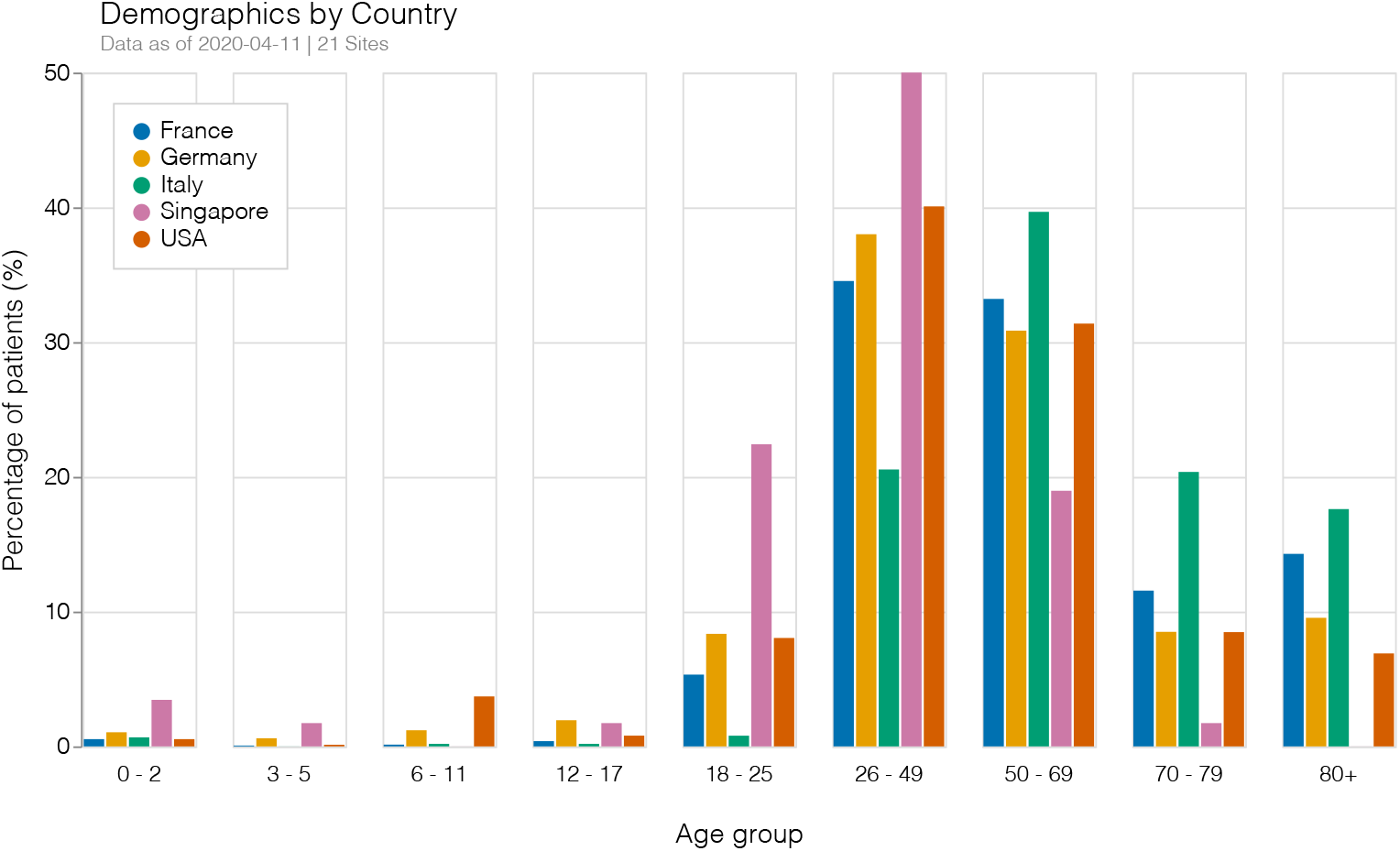
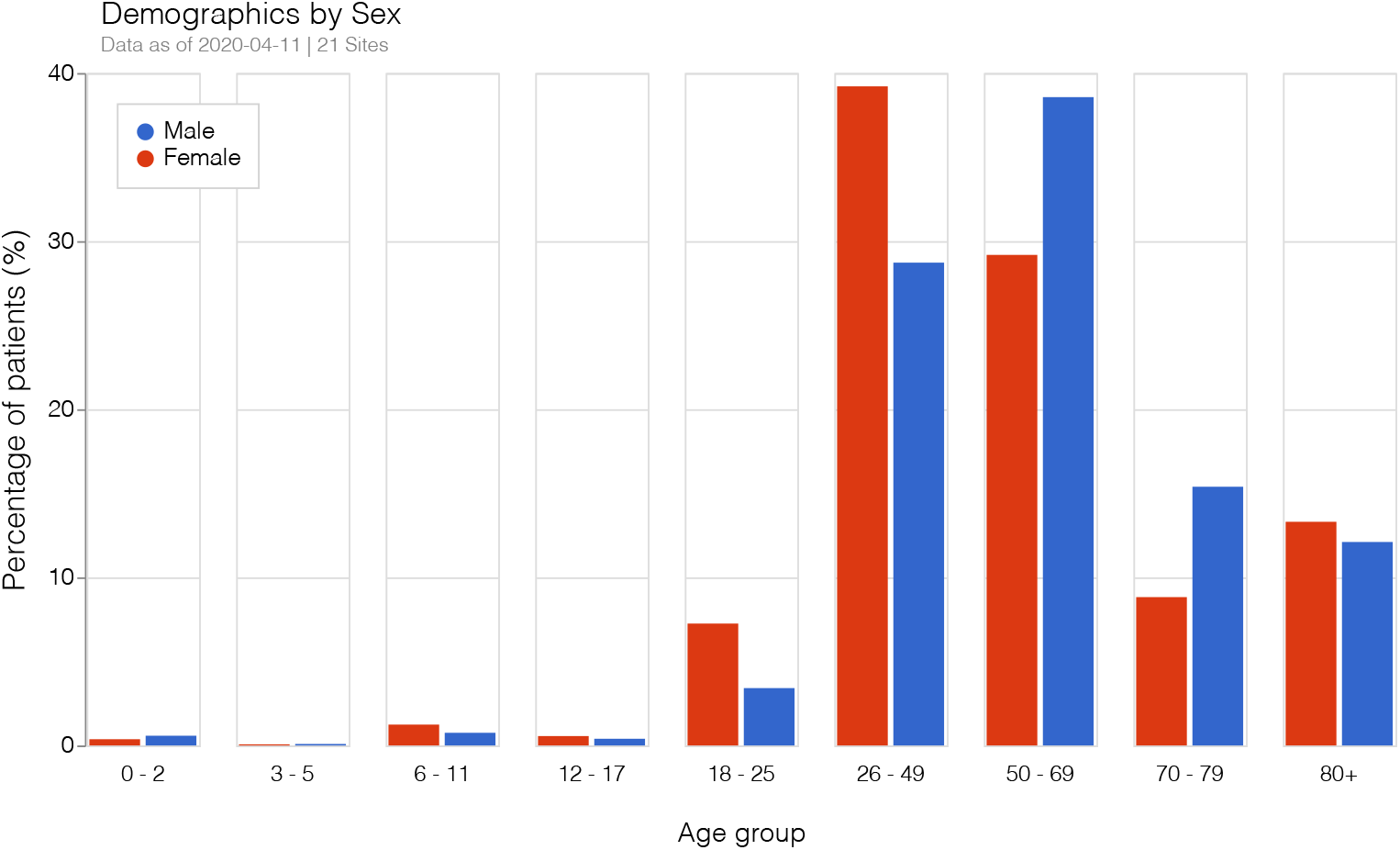
**(a)** Patients by country and age group. **(b)** Patients by sex and age group.

We were able to capture the total number of identified new cases by site and date. To normalize across sites with a small number of total cases, we generated the rate of growth of total admissions by country per day. In Figure 3, we compared those values with Johns Hopkins-curated data^24^ over time. Rates of growth as extracted from EHR data from our sites were similar to population-level findings during the month of March. Of note, national data from Singapore did not track with EHR numbers. Our one site in Singapore had a small number of patients and is less likely to be representative of all hospitals in the country.

**Figure 3.**
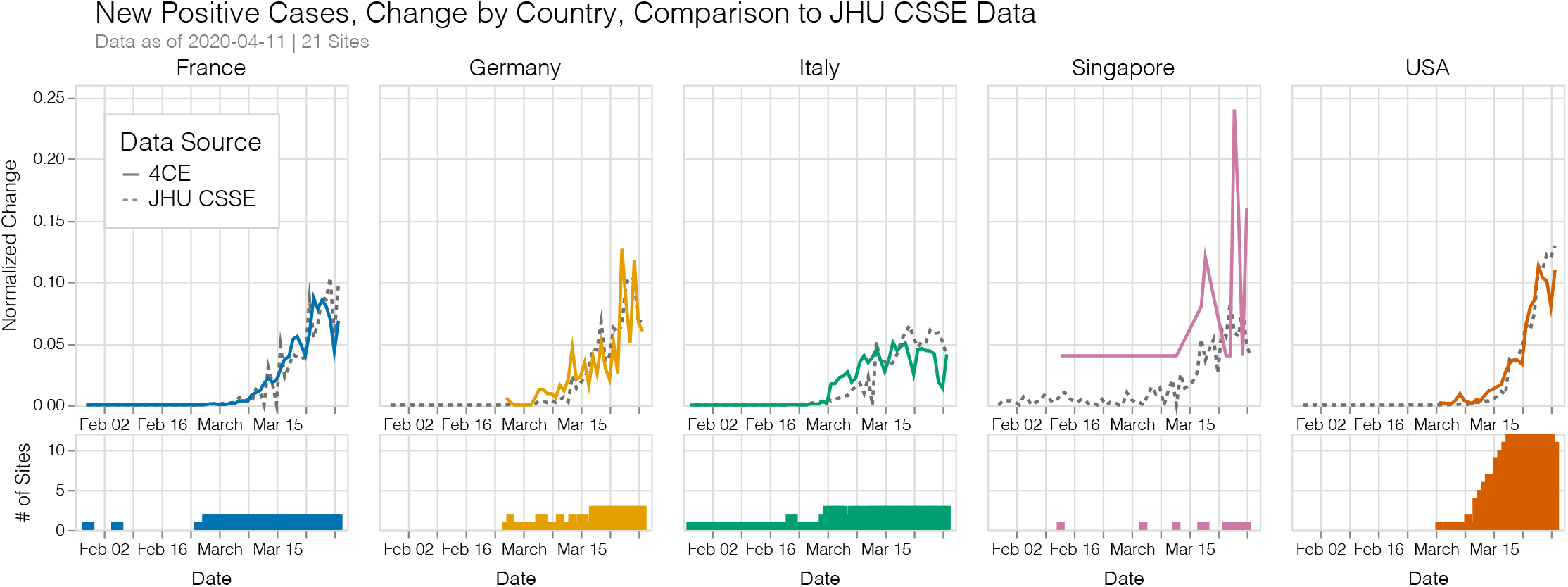
Normalized change (relative to previous day) of new cases reported by 4CE contributors compared to new cases collected by the Johns Hopkins Center for Systems Science and Engineering (JHU CSSE) by country over time. 4CE curves more closely resemble JHU CSSE numbers in relation to number of reported cases (e.g. newly reported cases on 3/31/2020: France - 1,124, Germany - 27, Italy - 106, Singapore - 2, USA - 394)

**Despite the limited amount of diagnosis code data submitted, consistent identified symptoms were recorded across institutions**. Most common codes involved respiratory symptoms and infections. Cough, dyspnea, and hyperpyrexia were also commonly identified. Although there were differences in the most common codes used by sites, symptoms were consistent with previous studies.^25,26^ Rates and types of presenting symptoms were similar in the pediatric population.

### Laboratory Value Trajectories

Our initial data extraction comprised 14 laboratory markers of cardiac, renal, hepatic, and immune dysfunction hat have been strongly associated with poor outcomes in COVID-19 patients in previous publications. Laboratory trajectories of each hospital at the population level are presented online at https://covidclinical.net. Given limitations of data harmonization and space, we focused on 5 laboratory trajectories that represented inflammatory, immune, hepatic, coagulation, and renal function. Trajectory data were remarkably consistent for most institutions at day 1 (day when biological test positive) with growing differences with continued hospitalization. Extensive data harmonization was performed, but we must emphasize that data from each day represented a potentially different population as patients were discharged, died, or laboratory studies were no longer performed. Data values from each hospital were an average of all studied patients a specified number of days after diagnosis.

**Laboratory values reflected relatively moderate disease severity on presentation**. Initial laboratory values were abnormal for all patients but were not indicative of organ failure. Major abnormal elevations were noted in C-reactive protein (CRP) and D-dimer on the day of diagnosis. As the number of days from diagnosis progressed, remaining patients who were not discharged or died had, on average, worse values. For nearly all 14 tests, trends toward progressively abnormal values were consistent with worsening disease as inpatient stays continued. Most importantly, the initial values and trajectories were highly consistent with previous findings in studies from China.^19,27^

**Creatinine**, a measure of renal function and the most commonly performed laboratory test in our data set, was divergent over time across sites. Rising creatinine would be consistent with an increased proportion of ill patients with significant acute kidney injury over time. Hospitals in Italy, in contrast, did not see a dramatic rise in creatinine in their hospitalized population, while the small percentage of French and German patients remaining in the hospital for two weeks had clear signs of acute kidney injury. This may represent many underlying differences including a high mortality near the beginning of the hospitalization at Italian hospitals, severe right time censoring of remaining patients, or a difference in practice.

**Total bilirubin**, a measure of conjugation and function by the liver, was initially normal across most sites and showed increases—consistent with other hepatic laboratory tests—among persistently hospitalized patients. The other hepatic laboratory measurements, ALT and AST, were divergent across institutions and showed a more significant perturbation (see https://covidclinical.net). Hepatic impairment was not present in most patients on presentation and total bilirubin was only mildly elevated with continued hospitalization.

On average, **white blood cell count** (WBC), a measure of immune response, was within normal limits on presentation. Patients who remained in the hospital and survived had increasing WBCs over time without severe leukocytosis.^27^ Lymphocyte and neutrophil count trajectories can be seen on the website. Procalcitonin and LDH were not commonly tested in the total patient population, but results are also online.

**C-reactive protein** (CRP), a measure of systemic inflammation, was notably elevated on presentation for all patients in the cohort with a very narrow confidence interval, consistent with previous findings.^19^ Although it is of unclear importance, populations of patients who remained in the hospital, survived, and had ongoing laboratory testing showed improvements over time. Interestingly, despite a decreasing trajectory during the first week, a mild leukocytosis is observed in counterbalance during the second week. The implication may be that CRP is not predictive of ongoing hospitalization or CRP is being checked for patient populations where the laboratory is more commonly improving.

**D-dimer**, an acute phase reactant and measure of coagulopathy, was elevated across institutions and countries at presentation. It rose consistently in all populations who continued to be hospitalized with the disease. This was consistent with multiple studies that showed a prothrombotic element to the disease. Most importantly, changes were consistent across all sites and highly abnormal.

**Figure 4.**
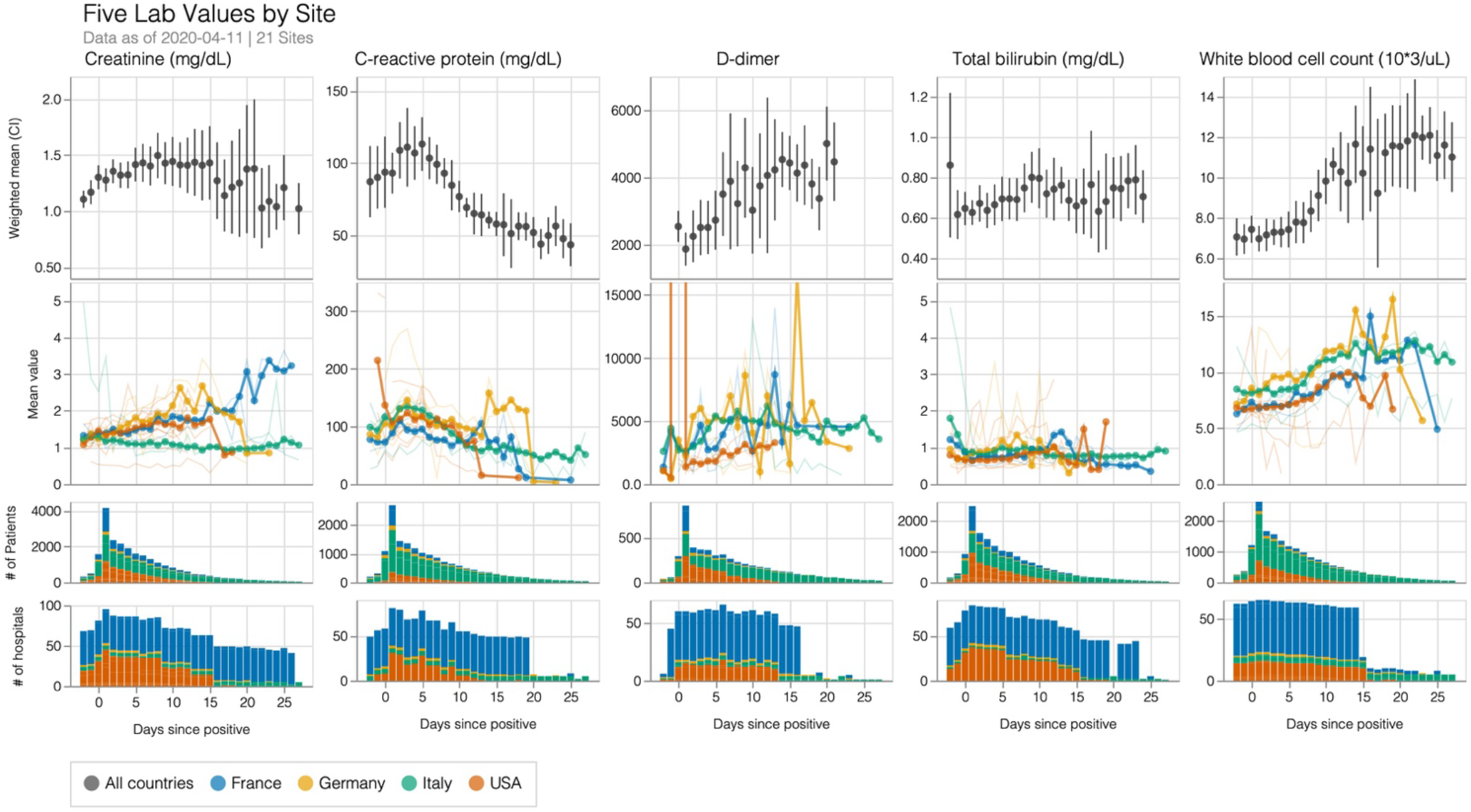
Laboratory tests representative of renal function (creatinine), systemic inflammation (C-reactive protein), coagulopathy (D-dimer), liver function (total bilirubin), and immune response (white blood cell count) visualized relative to date of diagnosis of COVID-19. The top row shows weighted means and 95% confidence intervals across all patients. The second row shows unweighted country-(thick lines) and site-level (thin lines) means. The third and fourth rows show the number of patients and sites, respectively, contributing laboratory tests of each type on a given day.

### Data Attrition

There was a large drop in the number of laboratory tests performed after the first day. Drop off in tests performed could be a result of death, length of stay, or change in frequency of data collection by the clinical team. From the maximum number of laboratory tests consistently checked on the first day after diagnosis, there was a rapid tapering in frequency of laboratory tests checked. These changes were particularly pronounced in Italy and France. We identified the number of days until the number of tests checked were 20% of their initial maximum value. Values for laboratory study for each day are presented on https://covidclinical.net. Results varied for each laboratory value and site. There was no obvious country level pattern. Given that several of these tests, such as creatinine, were commonly checked nearly every day in ill patients, the implication was that patients were censored from the laboratory results because of discharge or death or changing practice pattern. Thus, for the purposes of this paper, we focused on trends in creatinine. We normalized the number of tests performed by day to the total performed on day 1. We then looked at the day when the number of tests performed was 20% of the maximum number performed for each site. **For creatinine, for example, a drop-off in testing occurred between day 7 and 15 across institutions**. Most patients who survived were likely discharged within this time frame or managed with much less monitoring. Further results can be found online.

**Figure 5.**
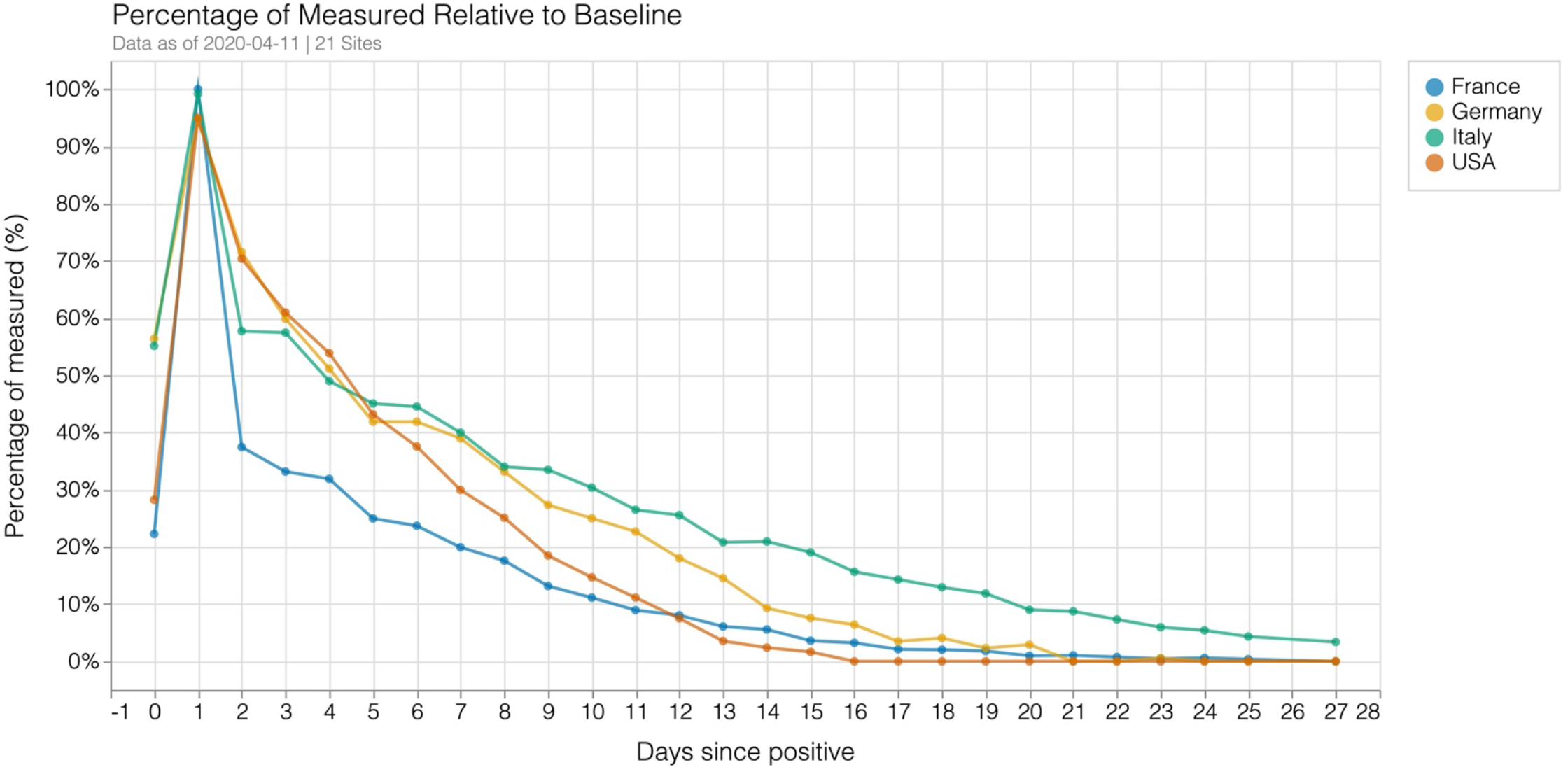
Drop-off in laboratory tests reported for creatinine relative to the day with the largest number of laboratory tests reported.

### Differences at Admission

There was greater between-hospital variation for laboratory test performance than between-country variation (Figure 6). At the time of diagnosis, there was significant variation between countries and between the hospitals in a specific country. There was no obvious signature presentation for a country for an individual laboratory value. For example, creatinine was a commonly performed laboratory study within a day of diagnosis. The overall standard deviation (SD) for creatinine values across countries was 1.47 while the SD within sites was 1.39. Standard deviation for countries was 1.64, 1.31, 1.13, and 1.62 within France, Germany, Italy, and the US, respectively. France was a special case as 39 hospitals were reported together by AP-HP and then compared with three hospitals in Bordeaux. This was an important finding that could suggest that laboratory values, as individual results, would not be able to fully explain the mortality differences between countries.

**Figure 6.**
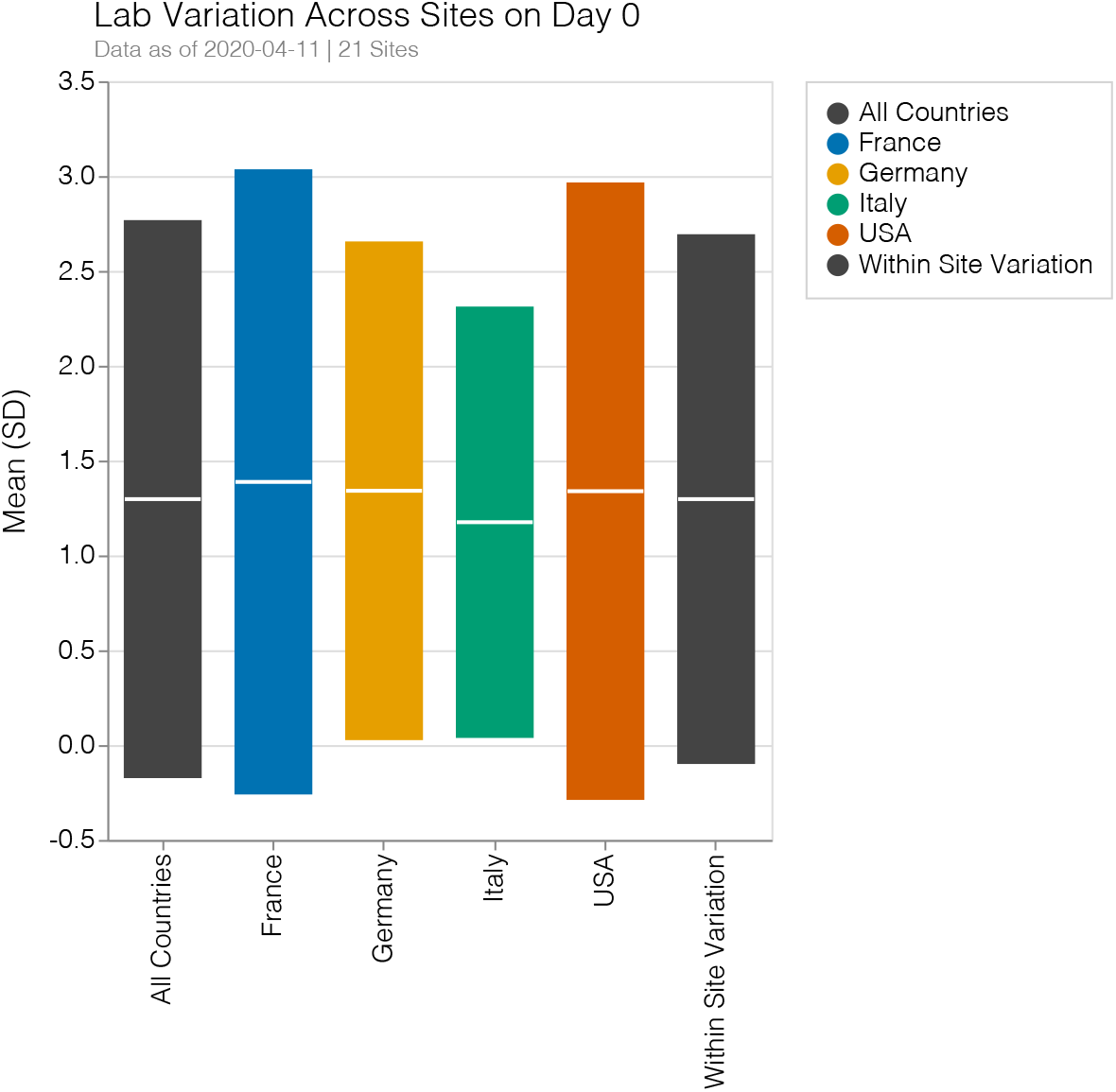
Laboratory variation across countries and within sites for creatinine test performed within a day of diagnosis. Values for mean value and standard deviation (SD) in creatinine level are shown. Large variations exist within sites and are often larger than the between country variation. The overall SD across countries was 1.47 while the SD within sites was 1.39. Standard deviation for countries was 1.64, 1.31, 1.13, and 1.62 within France, Germany, Italy, and the US, respectively.

## Discussion

A rapid mobilization of a multi-national consortium was able to harmonize and integrate data across 5 countries and 3 continents in order to begin to answer questions about comparative care of COVID-19 patients and opportunities for international learning. In just over 2 weeks, the group was able to define a question and data model, perform data extraction and harmonization, evaluate the data, and create a site for public evaluation of site level data. We aggregated electronic health record data from 96 hospitals, covering a total of 27,584 patients seen in these hospitals for COVID-19. In doing so, we relied upon prior investments made by various governments and institutions in turning the byproducts of clinical documentation into data useful for a variety of operational and scientific tasks, as documented in Table 1. Most importantly, at each site there were bioinformatics experts who understood both the technical characteristics of the data and their clinical relevance.

Using automated data extraction methods, we were able to show results consistent with country-level demographic and epidemiological differences identified in the literature. Rates of total case rise in our study was consistent with international tracking sites.^24^ Age breakdown, with Italian sites reporting a larger proportion of older patients, was also reflective of recent publicly available resources.^23^

We were able to show that laboratory trajectories across many hospitals could be collected and were concordant with findings from the literature. In truth, the findings generate more questions than they answer; the ability to see consistencies that spanned many countries indicated that the pathophysiology of this disease is shared across countries, and that demographics and care characteristics will have a significant effect on outcomes. As an example, the fall of CRP among those who continued to be hospitalized with a continued rise in d-dimer could suggest that d-dimer may be more closely related to persistent illness than CRP. The limits of our data collection method, where these results were not tied to the patient level and could not be associated across populations, highlights the need for caution with any conclusion related to changes in laboratory levels over time.

Perhaps most importantly, our study did not show a unique laboratory signature at the country level at the time of diagnosis. Researchers around the world have been closely following the rapid spread of COVID-19 and its high mortality rate in certain countries. One possible explanation would be that patients who presented to hospitals in Italy did so at a much more advanced stage of disease. Our results did not support this idea. There was as much in-hospital and between-hospital variation as between countries.

The average of laboratory values at presentation did not indicate major organ failure. This may be due to a larger proportion of healthier patients than those with advanced disease. Of course, respiratory failure could not be tracked within the limits of our data set.

There were both logistic and data interoperability lessons that were very important to the success of the project and will be critical for future efforts. Logistically, to maximize the timeliness of this consortium’s first collaboration around COVID-19, we deliberately aggregated the data to expedite the institutional review board (IRB) process at each institution for such data sharing. This constrained our analyses to count, rather than patient-level, data. While the latter would be optimal for deep analysis and identification of subtle patterns and perturbations of clinical courses, we felt that aggregated count data could provide valuable information on the clinical course even as we sought IRB permission for analyses at the patient level.

Regarding interoperability, large variations in units and data presentation required extensive data harmonization. The use of LOINC codes allowed for more rapid data extraction, but often institutions did not have internal mappings from their laboratory tests to LOINC codes. Manual interpretation of laboratory value descriptions were sometimes necessary. In future iterations, sites should perform unit conversion and ensure data consistency by presenting reference ranges and example data for a first-pass check of data at the site. Variations in ICD coding and inclusion made harmonization difficult. Frequencies of presenting codes were useful to show similar patterns to previous literature, but the current set of codes were too sparse for any further meaningful analysis. Future iterations of this project would encompass a much longer data capture timeline and would ensure comprehensive code collection across all sites.

In addition, data alignment by a metric that indicates clinical status is necessary to better establish outcomes. Using day of diagnosis as an alignment strategy did not allow for clear identification of causes for temporal patterns. Similarly, outcomes need to be selected that represent clinically meaningful endpoints secondary to this initial data alignment. One reason for this difficulty was that identification of level of care was not easily performed. Accordingly, it was not easy to follow patients in and out of ICUs at the site level and ICU data was not reliable.

Our group, the Consortium for Clinical Characterization of COVID-19 by EHR (4CE), is one of hundreds of efforts (some of which are listed at HealthIT.gov) that are working to aggregate and curate data to inform clinicians, scientists, policy makers and the general public. Additionally, networks of healthcare organizations such as the ACT network^28^ and PCORnet^29^ are working with federal authorities to obtain data-driven population-level insights. Similar initiatives are active in the other countries participating in 4CE, including the German Medical Informatics Initiative.^30^ Disease-specific and organ-specific COVID-19 research collectives are also assembling, including ones for cancers (https://ccc19.org), inflammatory bowel disease https://covidibd.org), and rheumatology,^31^ among many others. The World Health Organization maintains a directory of worldwide research efforts on COVID-19 including clinical data collection. ^32^ Finally, there are dozens of patient self-reporting apps with hundreds of thousands of users worldwide that provide perspectives on the clinical course of the infection outside hospitals.

There are a multitude of limitations to this study, not least of which is that it is observational and subject to a variety of biases. Perhaps the most severe is that study data is limited to those patients who were seen at or admitted to hospitals, due to severity of illness or other possibly biasing characteristics. Limitations also include heavy right censoring where patient absence can be due to death or discharge, variations in ICD annotations for conditions existing prior to the COVID-19-related admission, delays in updating billing codes or in uploading EHR data to the local analytic data repository. Furthermore, potentially confounding interactions between comorbidities, chronic diseases and their treatments and lifestyle or exposures were not taken into consideration. Again, because of these limitations we were careful to avoid making more than basic and descriptive conclusions. Over the coming weeks, we will work to quantify these biases and adjust for them, if we can. This will include adding data types as well as disaggregating the data to the patient level if and when permitted by IRBs. For the present, with the current limited knowledge of the clinical course of patients suffering from COVID-19, these results add to this small knowledge base. Our paper strikingly shows the power of harmonized data extraction from EHRs to rapidly study pandemics like COVID-19.

We invite others to join the 4CE consortium by sending a note to.

